# Comparison of influenza and COVID-19 hospitalizations in British Columbia, Canada: a population-based study

**DOI:** 10.1101/2022.08.26.22279284

**Authors:** Solmaz Setayeshgar, James Wilton, Hind Sbihi, Moe Zandy, Naveed Z Janjua, Alexandra Choi, Kate Smolina

## Abstract

**Objective:** To compare the population rate of COVID-19 and influenza hospitalizations by age, COVID-19 vaccine status and pandemic phase.

**Design:** Observational retrospective study

**Setting:** Residents of British Columbia (population 5.3 million), Canada

**Participants:** Hospitalized patients due to COVID-19 or historical influenza

**Main outcome measures:** This population based study in a setting with universal healthcare coverage, used COVID-19 case and hospital data for COVID-19 and influenza. Admissions were selected from March 2020 to February 2021 for the annual cohort and the first 8 weeks of 2022 for the peak cohort of COVID-19 (Omicron era). Influenza annual and peak cohorts were from three years with varying severity: 2009/10, 2015/16, and 2016/17. We estimated hospitalization rates per 100,000 population by age group.

**Results:** Similar to COVID-19 with median age 66 (Q1-Q3 44-80), influenza 2016/17 mostly affected older adults, with median age 78 (64-87). COVID-19 and influenza 2016/17 hospitalization rate by age group were “J” shaped. The rates for mostly unvaccinated COVID-19 patients in 2020/21 in the context of public health restrictions were significantly higher than influenza among individuals 30 to 69 years of age, and comparable to a severe influenza year (2016/17) among 70+. In early 2022 (Omicron peak), rates primarily due to COVID-19 among vaccinated adults were comparable with influenza 2016/17 in all age groups while rates among unvaccinated COVID-19 patients were still higher than influenza among 18+. In the pediatric population, COVID-19 hospitalization rates were similar to or lower than influenza.

**Conclusions:** Our paper highlighted the greater population-level impact of COVID-19 compared with influenza in terms of adult hospitalizations, especially among those unvaccinated. However, influenza had greater impact than COVID-19 among <18 regardless of vaccine status or the circulating variant.

## Introduction

On March 11, 2020, the World Health Organization (WHO) declared COVID-19 a global pandemic. Characteristics of this new respiratory disease were inevitably compared to seasonal influenza given both are largely respiratory diseases with similar symptom profiles albeit different mutation rates (although SARS-CoV-2 is being hypersequenced). However, studies suggested COVID-19 was associated with higher mortality, infectivity, along with a number of clinical differences.^1,2^ Each year in Canada (population 38.7 million) in the context of partially vaccinated population, it is estimated that influenza leads to approximately 12,000 hospitalizations^3^. In contrast, there have been close to 150,000 COVID-19 related hospitalizations in Canada during the course of two years of the pandemic.

There is a paucity of comparative studies on the respective morbidity and population burdens of COVID-19 and influenza epidemics, in particular by age and vaccination status.^1,2^ Instead, most comparative analyses have focused on severe outcomes among people who are already in hospital. Although data have shown higher disease severity for COVID-19 compared with seasonal influenza among in-hospital populations^2,4^, it is still unclear to what extent this is reflected at the population level or how rate of hospitalization for COVID-19 differs from influenza seasons with differing severities.

The epidemiology of COVID-19 is continually evolving as new variants emerge, new treatments are approved, and population immunity changes. The most recent variant (Omicron) in particular exhibits a different clinical and epidemiological profile.^5-8^ Before Omicron emerged, all homologous or heterologous mRNA and/or ChAdOx1 two-dose schedules were associated with ≥90% reduction in SARS-CoV-2 hospitalization risk for five to seven months.^9-11^ However, vaccine effectiveness against influenza hospitalization have been varied by age groups and ranged from negative values when vaccine components were antigenically distinct from the epidemic strain to ∼80% in case of high match.^12-15^ During the course of the COVID-19 pandemic, vaccination status of the Canadian population dramatically shifted from 100% unvaccinated in 2020 to over 90% of those aged 12 years and over having received at least one dose by the end of 2021. In contrast, historical coverage (2015 to 2019) for influenza vaccine in Canada has been about 33% of adults overall, ranging between 20% in 18-34 year olds to 60% in 65+ year olds.^16^

As jurisdictions transition to COVID-19 endemicity with likely co-circulation of influenza, comparing COVID-19 hospitalization rates to historical influenza seasons is vital for informing healthcare planning. Moreover, during the pandemic public health measures have slowed transmission but created unintended consequences. Going forward, understanding the relative morbidity and mortality of COVID-19 compared to more familiar pathogens will be important to policymakers contemplating a sustainable approach. In this study, we compared the population rate of COVID-19 and influenza hospitalizations by age, COVID-19 vaccine status and pandemic phase among residents of British Columbia, Canada. Our analysis captures the most recent Omicron wave and adjusts hospitalization estimates to only include those admitted primarily for COVID-19.

## Method

### Data source

This study used data from the British Columbia COVID-19 Cohort (BCC19C), a public health surveillance platform integrating COVID-19 datasets (testing, cases, hospitalizations, vaccinations) with other administrative data holdings for the BC population (see Table S1 for list of linked datasets). The BCC19C was established as a public health surveillance system and is maintained through operational support from Data Analytics, Reporting and Evaluation (DARE), and BC Centre for Disease Control (BCCDC) at the Provincial Health Services Authority.

### Study Design, Setting, and Participants

British Columbia (BC) is the westernmost province of Canada, with a population of 5.26 million individuals and universal healthcare coverage. We created retrospective cohorts of inpatients with at least an overnight hospital stay due to COVID-19 or influenza. Each cohort was divided into annual (12 months) and seasonal peak (eight weeks) cohorts based on date of hospitalization. In the annual cohort analysis, we compared hospitalization rates (COVID-19 vs influenza) when the majority of the BC population were unvaccinated during the period from March 2020 to February 2021 (by the end of February 2021, fewer than 10% of BC residents in each age group had received at least one dose, the majority occurring towards the end of the month). In the seasonal peak comparison, we included the first Omicron wave in BC in January and February 2022 and stratified COVID-19 rates by vaccination status and whether hospitalization was primarily for COVID-19 or not (Figure S1). Data on influenza vaccination was not available in order to stratify influenza hospitalizations by vaccination status.

#### Influenza hospitalizations

Influenza hospitalizations were identified from the Discharge Abstract Database (DAD).^17^ DAD data captures admissions, discharges, transfers, and deaths occurring in acute care hospitals in BC. International Classification of Diseases codes are used for diagnostic coding.

In the absence of harmonized laboratory testing information for the whole province to confirm presence of influenza virus, we applied a validated algorithm (FLU2) using International Classification of Diseases 10^th^ edition (ICD-10).^18^ The FLU2 ICD-10 codes included J09, J10.0, J10.1, J10.8, J11.0, J11.1, and J11.8. We further validated it in BC setting using data for one region in BC for which influenza test results were available (see Sensitivity Analysis).

Influenza seasons from 2020-2022 were atypical. In order to compare COVID-19 hospitalizations to more typical influenza seasons, we selected three different seasons of varying severity and circulating strains to ensure a more balanced comparison. For consistency in peak definitions with the COVID-19 (Omicron) peak, we chose comparator influenza seasons that only had a single peak in order to be able to capture a continuous 8-week period of highest levels of hospitalizations in a season. Influenza 2009/10 was H1N1 pandemic causing severe disease in younger people, but was less potent as the elderly had baseline immunity. Influenza 2015/16 saw mild influenza A (H1N1) and B (Victoria) activity mainly affecting children whereas influenza 2016/17 saw severe influenza A (H3N2) activity mainly affecting older adults. Seasonal peak cohorts included a data-driven eight-week peak of hospital admissions: October 28, 2009 to November 28, 2009 (influenza 2009/10); January 24, 2016 to March 19, 2016 (influenza 2015/16); and December 18, 2016 to February 11, 2017 (influenza 2016/17). Annual (12 months) cohorts included hospitalizations between September and August comprising the above influenza seasons (2009/10, 2015/16, 2016/17).

#### COVID-19 hospitalizations

Our study extracted COVID-19 hospitalization data from Integrated COVID-19 laboratory dataset^19^ that included COVID-19 case surveillance data^20^ and Provincial COVID-19 Monitoring Solution (PCMS).^21^ We included all patients hospitalized for COVID-19 up to 14 days after or 2 days prior to specimen collection date in order to exclude hospitalization due to reasons other than COVID-19. We did not use DAD data for COVID-19 hospitalization due to 5-6 month lag in DAD data updates.

To capture a time period when most of the BC population was unvaccinated, the annual cohort included all patients hospitalized for COVID-19 between March 2020 and February 2021 during which the first Variant of Concern, Alpha, emerged and less than 10% in each age group had received at least one dose. For the seasonal peak cohort, we included individuals admitted to hospital during the time where the rate of COVID-19 hospitalizations was the highest and when fewer restrictions were in place (the first 8 weeks of 2022 when Omicron was dominant).

To mitigate selection bias, we attempted to account for hospitalizations that were not primarily due to COVID-19 (i.e., hospitalizations among COVID-19 positive patients hospitalized for non COVID-19 reasons whose infection was likely identified through routine screening) in the seasonal analysis. According to an internal chart review of 600 patients hospitalized in December 2021 and January 2022 in BC, less than half of COVID-19 related hospitalizations during Omicron era were primarily due to COVID-19, with differences by age.^22^ Leveraging these data and other data sources,^23^ we stratified aggregate rates into 1) estimated proportion admitted *primarily due to COVID-19* (severe or moderate symptoms and COVID-19 was the most responsible diagnosis for admission) and, 2) estimated proportion admitted *primarily for other reasons* (no diagnosis at the time of admission and mild symptoms). The following estimates for proportion of hospitalizations likely primarily due to COVID-19 were: unvaccinated <50 years old (50%), unvaccinated ≥50 years old (70%), fully vaccinated <50 years old (20%) and fully vaccinated ≥50 years old (40%).

### Variables

Variables of interests are presented in Table S2. Hospital admission was stratified by age groups, sex, health authority of residence, and vaccination status where available. Missing information was categorized as “Unknown”.

### Statistical Analysis

#### Descriptive

We summarized patients’ demographic information by frequencies (percentages) and by median (Q1-Q3) for categorical and continuous variables, respectively. We compared length of stay (LOS) for COVID-19 and influenza related hospitalization, using Kruskal–Wallis one-way analysis of variance for all patients and by age group.

We compared influenza and COVID-19-related hospitalization rates per 100,000 population for both the annual and seasonal cohorts. Rates were calculated by dividing the total number of hospitalizations by annually updated population denominators obtained from census-based BC Stats Population Estimates and projections^24^ or from provincial immunization registry data for COVID-19 vaccinated and unvaccinated denominators.

#### Poisson regression (annual analysis)

We applied a Poisson regression model for the annual analysis and compared hospitalization rate of influenza in 2009/10, 2015/16, and 2016/17 with COVID-19 while adjusting for age, sex, and health authority to calculate incidence rate ratios. In addition, we analyzed age-specific incidence rate ratios and adjusted for sex and health authority. Due to the large uncertainty in the estimates of hospitalization primarily due to COVID-19 in the younger age groups, the adjusted rate ratio for the peak analysis were not computed..

#### Sensitivity analysis and validation

Influenza hospitalizations was identified using a different approach and compared to our primary analysis. Here, influenza testing data was linked to DAD to identify all hospitalizations (regardless of ICD-10 codes) within three days of a laboratory-confirmed influenza diagnosis. We limited this sensitivity analysis to one of the most populous regional health authority, Vancouver Coastal Health (VCH population 1.26 million, or 24% of BC population), for which influenza testing data were complete (data for other regions were not complete – precluding the use of this approach for the primary analysis). We calculated sensitivity, specificity, positive predictive value (PPV) and negative predictive value (NPV) of the ICD-10 FLU2 approach used in our primary analysis, using hospitalizations with a confirmed positive influenza test as the gold standard.

Analyses were conduced using SAS version 9.4, and graphs and rate ratio calculations were produced using R Studio version 3.6.2.

#### Patient and public involvement

Discussions and debates between members of the public, patients, public servants, and decision makers on how COVID-19 compares with influenza to inform preparations for the time of their likely co-circulation in the fall 2022 have inspired this study. No patients were directly involved in the production of this article.

## Results

Our seasonal peak analysis during Omicron era and in the context of mostly vaccinated population (>90%) included 4,340 COVID-19 hospitalizations, of which ∼1,870 (43%) were estimated to be primarily due to COVID-19. Numbers of influenza hospitalizations were lower: 1,272 (2009/10), 538 (2015/16) and 1,319 (2016/17) (Table 1). Similarly, the annual analysis during first year of the pandemic in the context of mostly unvaccinated population included 3,398 COVID-19 hospitalizations and the number of influenza hospitalizations for 2009/10, 2015/16, and 2016/17 were 1,483, 1,032, and 1,979 respectively (Table S3).

**Table 1:** Demographic characteristics of hospitalized patients testing positive for COVID-19 during the first 8 weeks of 2022 (Omicron era) and for seasonal influenza, the 8-week peak of 2009/10 (H1N1 pandemic), 2015/16 (severe for children), and 2016/17 (severe for adults) in British Columbia, Canada

| Characteristics | COVID-19, total<br>(n=4,340) | Primarily for COVID-19*<br>(n=1,870) | Influenza 2009/10<br>(n=1,272) | Influenza 2015/16<br>(n=538) | Influenza 2016/17<br>(n=1,319) |
| --- | --- | --- | --- | --- | --- |
|  | n (%) | n (%) | n (%) | n (%) | n (%) |
| **Sex** |  |  |  |  |  |
| Female | 1975 (46) | 801 (43) | 675 (53) | 269 (50) | 695 (53) |
| Male | 2330 (54) | 1053 (56) | 597 (47) | 269 (50) | 624 (47) |
| Unknown | 35 (1) | 15 (1) | 0 (0) | 0 (0) | 0 (0) |
| **Age, years** |  |  |  |  |  |
| Median (Q1-Q3) | 66 (44-80) | NA | 39 (15-55) | 56 (24-72) | 78 (64-87) |
| **Age category, years** |  |  |  |  |  |
| 0-4 | 132 (3) | 66 (4) | 175 (14) | 70 (13) | 45 (3) |
| 5-11 | 45 (1) | 19 (1) | 117 (9) | 40 (7) | 11 (1) |
| 12-17 | 66 (2) | 17 (1) | 54 (4) | 16 (3) | 13 (1) |
| 18-29 | 280 (6) | 82 (4) | 167 (13) | 23 (4) | 21 (2) |
| 30-39 | 419 (10) | 116 (6) | 124 (10) | 44 (8) | 45 (3) |
| 40-49 | 340 (8) | 97 (5) | 190 (15) | 36 (7) | 44 (3) |
| 50-59 | 429 (10) | 206 (11) | 212 (17) | 71 (13) | 100 (8) |
| 60-69 | 724 (17) | 362 (19) | 105 (8) | 83 (15) | 164 (12) |
| 70+ | 1905 (44) | 905 (48) | 128 (10) | 155 (29) | 876 (66) |
| **Health authority** |  |  |  |  |  |
| Fraser | 2027 (47) | 846 (45) | 406 (32) | 128 (24) | 306 (23) |
| Vancouver Coastal | 853 (20) | 362 (19) | 241 (19) | 167 (31) | 406 (31) |
| Vancouver Island | 406 (9) | 175 (9) | 177 (14) | 129 (24) | 416 (32) |
| Interior | 809 (19) | 376 (20) | 307 (24) | 67 (12) | 154 (12) |
| Northern | 245 (6) | 112 (6) | 128 (10) | 40 (7) | 22 (2) |
| Unknown | 0 (0) | 0 (0) | 13 (1) | 7 (1) | 15 (1) |
| **Vaccination** |  |  |  |  |  |
| Unvaccinated | 1300 (30) | 817 (44) | NA | NA | NA |
| Vaccinated, 1 dose | 182 (4) | 58 (3) | NA | NA | NA |
| Vaccinated, 2+ doses | 2858 (66) | 995 (53) | NA | NA | NA |

| Characteristics | COVID-19,<br>total<br>(n=4,340) | Primarily for<br>COVID-19*<br>(n=1,870) | Influenza<br>2009/10<br>(n=1,272) | Influenza<br>2015/16<br>(n=538) | Influenza<br>2016/17<br>(n=1,319) |
| --- | --- | --- | --- | --- | --- |
| Abbreviation: COVID-19=Coronavirus Disease 2019, n=number, NA=Not Applicable, Q=Quartile<br>*We applied the following estimates for proportion of hospitalizations likely primarily due to COVID-19: unvaccinated <50 years old (50%), unvaccinated ≥50 years old (70%), vaccinated <50 years old (20%) and vaccinated ≥50 years old (40%). Numbers were rounded to zero decimal point.<br>Note: For the peak COVID-19 cohort we included all patients hospitalized for COVID-19 up to 14 days after or 2 days prior to specimen collection date during the first 8 weeks of 2022 when Omicron was dominant and >90% of adults in British Columbia were vaccinated with at least 2 doses. We selected the 8-week peak of three seasons with distinct severity for influenza: 2009/10=H1N1 pandemic, 2015/16=mild with higher severity in children, 2016/17=severe with higher severity in adults. |  |  |  |  |  |

### Patient characteristics

Characteristics of hospitalized patients are presented in Table 1 (seasonal peak analysis) and Supplementary Table S3 (annual analysis). In both cohorts, a higher proportion of COVID-19 hospitalizations were male relative to influenza. The age distribution differed by influenza seasons. Similar to COVID-19 with median age 66 (Q1, Q3 44, 80), influenza 2016/17 mostly affected older adults, with median age 78 (64, 87), while influenza 2015/16, and especially influenza 2009/10 (H1N1), affected younger individuals (median (Q1, Q3) 56 (24, 72) and 39 (15, 55), respectively) (Table 1). Similar findings were observed for the annual analysis (Table S3). Among children <18, the highest proportion of hospitalizations were among 0-4 year olds for both COVID-19 and influenza while for adults >50, the proportion increased by age for all seasons studied except for 2009/10, likely due to previous exposure to H1N1 among older adults.

### Hospitalization rates

#### Annual Analysis

In the annual analysis (first year of the pandemic; largely unvaccinated population; restrictions in place), the COVID-19 hospitalization rate by age group was “J” shaped, similar to influenza 2016/17 (partly vaccinated population; no restrictions in place) (Figure 1). Among individuals 0 to 17 years of age, COVID-19 hospitalizations were significantly lower than all annual cohorts of influenza. However, the COVID-19 hospitalization rates were significantly higher than those for all influenza years studied among individuals 30 to 69 years of age, and comparable to a severe influenza year (2016/17) among 70+ (Figure 1).

**Figure 1:**
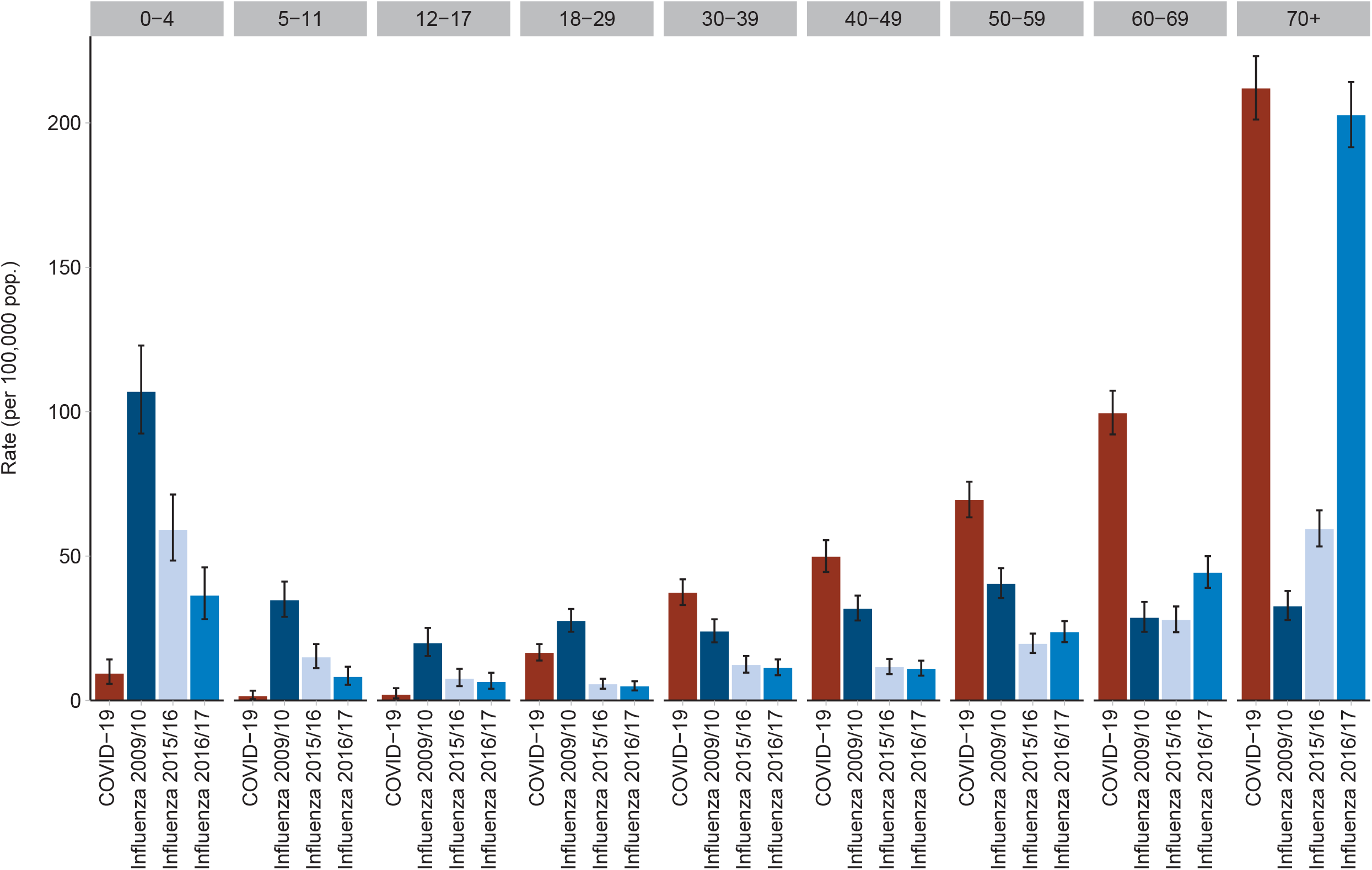
Annual hospitalization rates for patients testing positive for COVID-19 (2020/21) compared with patients hospitalized for influenza 2009/10 (H1N1 pandemic), 2015/16 (severe for children), and 2016/17 (severe for adults), by age group, British Columbia, Canada Note: For the annual COVID-19 cohort we included all patients hospitalized for COVID-19 up to 14 days after or 2 days prior to specimen collection date from March 2020 to February 2021. For influenza, we selected three 12-month periods from September to August with distinct severity: 2009/10=H1N1 pandemic, 2015/16=mild influenza with higher severity in children, 2016/17=severe influenza with higher severity in adults.

#### Seasonal Peak Analysis

The seasonal peak analysis (Omicron era; largely vaccinated population; fewer restrictions) revealed similar trends. COVID-19 hospitalizations again displayed a “J” shaped trend, with the rate of hospitalization for COVID-19 increasing by age in adults (Figure 2). The estimated rate of hospitalization primarily for COVID-19 per 100,000 population (30 (95% CI 23 to 38)) in unvaccinated 0-4 year olds was about four times higher compared with unvaccinated older children 5-11 (8 (5 to 13)) and 1.5 times higher than 12-17 year olds (20 (8 to 41)).

**Figure 2:**
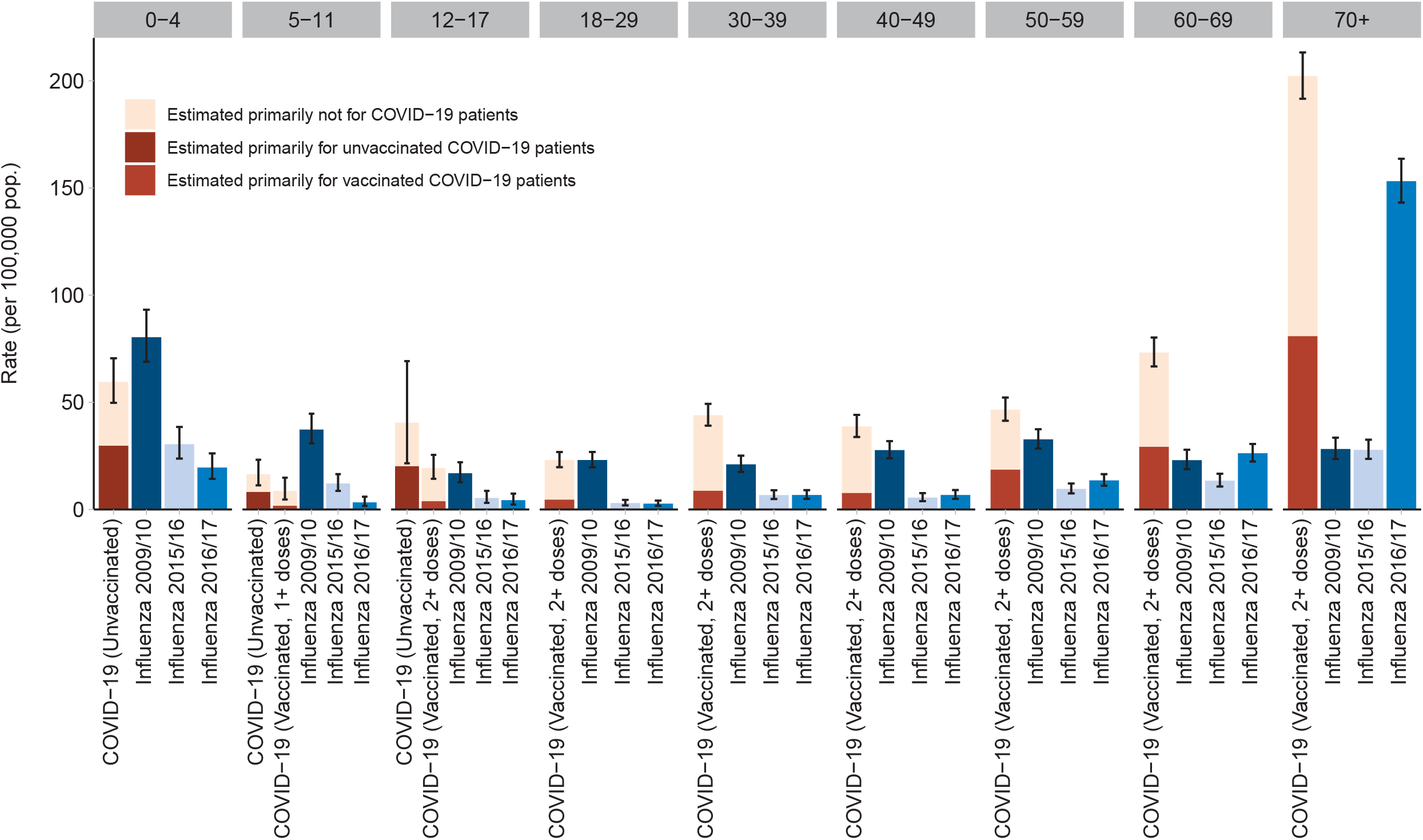
Hospitalization rates in patients testing positive for COVID-19 (excluding unvaccinated 18+) during the peak of Omicron variant (week 1-8 of 2022) compared with hospitalized patients for influenza in the peak of 2009/10 (H1N1 pandemic), 2015/16 (severe for children), and 2016/17 (severe for adults) season, by age group and COVID-19 vaccine status, British Columbia, Canada Note: For the peak COVID-19 cohort we included all patients hospitalized for COVID-19 up to 14 days after or 2 days prior to specimen collection date during the first 8 weeks of 2022 when Omicron was dominant and >90% of adults in British Columbia were vaccinated with at least 2 doses. We selected the 8-week peak of three seasons with distinct severity for influenza: 2009/10=H1N1 pandemic, 2015/16=mild with higher severity in children, 2016/17=severe with higher severity in adults. We applied the following estimates for proportion of hospitalizations likely primarily due to COVID-19: unvaccinated <50 years old (50%), unvaccinated >=50 years old (70%), vaccinated <50 years old (20%) and vaccinated >=50 years old (40%).

Rates of hospitalization primarily due to COVID-19 among vaccinated adults were comparable with a severe influenza year (2016/17) for all age groups except those 70+ (higher for influenza). Rates for 12-49 year olds were comparable to a milder influenza year (2015/16). Hospitalizations rates for pandemic influenza 2009/10 were higher than those primarily due to COVID-19 among 0-59 years of age and lower among 60+, again likely attributable to prior exposure (Figure 2). In contrast, rates among those unvaccinated for COVID-19 remained higher than influenza for individuals aged 18+ years (Figure S2). The same was not true for unvaccinated individuals 0-17 years old, for whom hospitalization rates for COVID-19 remained lower or comparable to influenza.

### Rate ratio

Table 2 shows adjusted incidence rate ratios (aIRR) comparing the hospitalization rate of influenza (partially vaccinated population, no restrictions) to COVID-19 in 2020/21 (largely unvaccinated population, restrictions in place) for the annual analysis. The overall model showed that overall rate of hospitalization for influenza was between 50% and 70% lower than COVID-19 after adjusting for age group, sex, and health authority. When stratified by age group, the rate of influenza hospitalization was highest for the pediatric population.

**Table 2:** Annual analysis: Rate ratio of hospitalization for influenza in 2009/10 (H1N1 pandemic), 2015/16 (severe for children), and 2016/17 (severe for adults) compared to hospitalized patients testing positive for COVID-19 during a year in 2020/21 by age groups, British Columbia, Canada

| **Population<sup>§</sup>** | **Rate Ratio (95% CI)** |
| --- | --- |
| **All age groups** |  |
| Influenza 2009/10 | 0.5 (0.4-0.7) |
| Influenza 2015/16 | 0.3 (0.3-0.4) |
| Influenza 2016/17 | 0.6 (0.4-0.8) |
| **0-4 y** |  |
| Influenza 2009/10 | 6.1 (4.0-9.4) |
| Influenza 2015/16 | 3.4 (2.1-5.3) |
| Influenza 2016/17 | 2.3 (1.4-3.7) |
| **5-11 y** |  |
| Influenza 2009/10 | 12.8 (10.0-16.4) |
| Influenza 2015/16 | 5.6 (4.1-7.5) |
| Influenza 2016/17 | 3.0 (2.2-4.2) |
| **12-17 y** |  |
| Influenza 2009/10 | 6.8 (4.6-10.1) |
| Influenza 2015/16 | 2.9 (1.9-4.4) |
| Influenza 2016/17 | 2.7 (1.6-4.6) |
| **18-29 y** |  |
| Influenza 2009/10 | 1.7 (1.3-2.2) |
| Influenza 2015/16 | 0.4 (0.2-0.5) |
| Influenza 2016/17 | 0.3 (0.2-0.4) |
| **30-39 y** |  |
| Influenza 2009/10 | 0.6 (0.4-0.9) |
| Influenza 2015/16 | 0.3 (0.2-0.5) |
| Influenza 2016/17 | 0.3 (0.2-0.4) |
| **40-49 y** |  |
| Influenza 2009/10 | 0.6 (0.4-0.9) |
| Influenza 2015/16 | 0.2 (0.1-0.3) |
| Influenza 2016/17 | 0.2 (0.1-0.3) |
| **50-59 y** |  |
| Influenza 2009/10 | 0.6 (0.4-0.8) |
| Influenza 2015/16 | 0.3 (0.2-0.4) |
| Influenza 2016/17 | 0.3 (0.2-0.5) |
| **60-69 y** |  |
| Influenza 2009/10 | 0.3 (0.2-0.4) |
| Influenza 2015/16 | 0.3 (0.2-0.4) |
| Influenza 2016/17 | 0.4 (0.3-0.6) |
| **70+ y** |  |
| Influenza 2009/10 | 0.2 (0.1-0.2) |
| Influenza 2015/16 | 0.3 (0.2-0.4) |
| Influenza 2016/17 | 0.9 (0.6-1.5) |

| Population<sup>§</sup> | Rate Ratio (95% CI) |
| --- | --- |
| Abbreviation: COVID-19=Coronavirus Disease 2019, CI=Confidence Interval<br>Note: All rate ratios, except influenza 2016/17 among 70+ (p-value=0.179), were statistically significant with p-value <0.001. For the annual COVID-19 cohort we included all patients hospitalized for COVID-19 up to 14 days after or 2 days prior to specimen collection date from March 2020 to February 2021. For influenza, we selected three 12-month periods from September to August with distinct severity: 2009/10=H1N1 pandemic, 2015/16=mild influenza with higher severity in children, 2016/17=severe influenza with higher severity in adults<br><sup>§</sup>Reference group was rate for COVID-19. The 'all age group' Poisson model was adjusted for age categories, sex, and health authorities. The age specific models were adjusted for sex and health authorities. |  |

### Length of Stay in the hospital

In the seasonal peak analysis, the overall median for length of stay (LOS) in the hospital due to COVID-19 was 6 days (Q1, Q3 3, 13), which was similar to a severe influenza year (2016/17) (6 (3, 12)) (Table 3). In age stratified analyses, median LOS was significantly higher for COVID-19 patients (vs. influenza) among 60+ years of age (Table 3). In the annual analysis, the LOS for COVID-19 patients was significantly higher than influenza among 18+ years of age but not in the pediatric population (Table S4).

**Table 3:** Median of length of stay (days) in hospital among patients testing positive for COVID-19 during the first 8 weeks of 2022 (Omicron era) and among patients hospitalized for seasonal influenza during the 8-week peak of 2009/10 (H1N1 pandemic), 2015/16 (severe for children), and 2016/17 (severe for adults) in British Columbia, Canada

| Age group, years | COVID_19<br>(n=4316<sup>§</sup>) | Influenza<br>2009/10<br>(n=1272) | Influenza<br>2015/16<br>(n=538) | Influenza<br>2016/17<br>(n=1319) | P-value<sup>#</sup> |
| --- | --- | --- | --- | --- | --- |
| **Total** |  |  |  |  |  |
| Median (Q1-Q3) | 6 (3-13) | 3 (2-7) | 5 (2-9) | 6 (3-12) | <0.001 |
| **0-4** |  |  |  |  |  |
| Median (Q1-Q3) | 2 (1-3) | 2 (1-4) | 3 (2-5) | 3 (1-6) | 0.001 |
| **5-11** |  |  |  |  |  |
| Median (Q1-Q3) | 2 (2-5) | 2 (2-5) | 3 (2-6) | 2 (1-3) | 0.419 |
| **12-17** |  |  |  |  |  |
| Median (Q1-Q3) | 2 (2-5) | 2.5 (2-5) | 1.5 (1-4) | 3 (1-3) | 0.375 |
| **18-29** |  |  |  |  |  |
| Median (Q1-Q3) | 2 (2-5) | 2 (1-4) | 3 (2-5) | 3 (1-3) | 0.575 |
| **30-39** |  |  |  |  |  |
| Median (Q1-Q3) | 3 (2-5) | 3 (1-5) | 3 (2-9) | 3 (2-5) | 0.175 |
| **40-49** |  |  |  |  |  |
| Median (Q1-Q3) | 4 (2-8) | 4 (2-7) | 4 (2-6.5) | 4 (2-7.5) | 0.781 |
| **50-59** |  |  |  |  |  |
| Median (Q1-Q3) | 6 (3-11) | 5 (2-8) | 6 (3-16) | 3 (2-6) | <0.001 |
| **60-69** |  |  |  |  |  |
| Median (Q1-Q3) | 7 (4-15) | 5 (3-9) | 5 (3-8) | 4 (3-9) | <0.001 |
| **70+** |  |  |  |  |  |
| Median (Q1-Q3) | 8 (4-17) | 6 (3.5-11) | 7 (4-12) | 7 (4-14) | <0.001 |

Abbreviation: COVID-19=Coronavirus Disease 2019, n=number, Q=Quartile
Note: For the peak COVID-19 cohort we included all patients hospitalized for COVID-19 up to 14 days after or 2 days prior to specimen collection date during the first 8 weeks of 2022 when Omicron was dominant and >90% of adults in British Columbia were vaccinated with at least 2 doses. We selected the 8-week peak of three seasons with distinct severity for influenza: 2009/10=H1N1 pandemic, 2015/16=mild with higher severity in children, 2016/17=severe with higher severity in adults.
<sup>§</sup>The numbers are based on the availability of both admission and discharge date. n=24 patients with COVID-19 did not have discharge date, thus, excluded.
<sup>#</sup>Kruskal–Wallis one-way analysis of variance

### Sensitivity analysis and method validation

When comparing case ascertainment using lab-based data to the primary analysis whereby we relied on administrative health data (Figure S3), we demonstrated overall good sensitivity across all flu seasons with 88% (95% CI: 83, 93), 71% (65, 77), 75% (71, 79) for 2009/10, 2015/16, and 2016/17, respectively (Table S5). Further, overall population level rate estimates were comparable using ICD codes versus positive lab-confirmed influenza tests in all influenza years, with overlapping CIs (Figure S3).

## Discussion

We created retrospective, population-based cohorts of COVID-19 and influenza hospitalizations during different pandemic phases. Among adults, our analysis showed that pre-Omicron, in the context of a largely unvaccinated population and stronger public health measures, COVID-19 related hospitalization rates were significantly higher than historical influenza rates. However, in 2022 with a largely vaccinated population despite fewer restrictions, hospitalization rates for COVID-19 were similar to historical influenza rates. Unvaccinated adults (18+) during this period remained at risk with higher COVID-19 hospitalization rates. In contrast, among those 0-17 COVID-19 hospitalization rates in the first two years of the pandemic were comparable or lower than those for influenza regardless of vaccination status or circulating variant. This is particularly notable given that seroprevalence studies in BC and Quebec (January-February 2022) showed a large proportion of previously infected children under the age of five at a point when few were vaccinated.^25^ This should be interpreted in the context of contemporaneous public health measures potentially lowering contact rates.

Our findings are similar to other studies reporting higher rates of hospitalization for COVID-19 compared to influenza in unvaccinated populations in the early phases of the pandemic.^2,26^ In a French study, there were about 2 times the number of COVID-19 hospitalizations in a 2-month period (89,530; March 1^st^ to April 30^th^, 2020) compared to a 3-month period for seasonal influenza (N=45,819; Dec 1^st^ 2018 to Feb 28^th^, 2019).^2^ In an analysis comparing COVID-19 in a two month period in 2020 to the previous five influenza seasons (8 months each) at a large hospital in Boston, there were 582 COVID-19 hospitalizations compared to an average of 210 influenza admissions.^26^ Importantly, the relative difference between COVID-19 and influenza is likely underestimated in these comparisons due to strong public health measures implemented during COVID-19, high influenza vaccination coverage, and prior influenza infection conferring protection from severe disease. In contrast to studies examining unvaccinated individuals, we found that COVID-19 hospitalization rates in fully vaccinated individuals were generally comparable to influenza rates in individuals aged 12-69, particularly when we excluded hospitalizations not primarily due to COVID-19. Both COVID-19 and influenza 2016/17 had “J” shaped age-specific hospitalization rate curves, with higher rates at the extremes of age. Similar to the French study, the rate of hospitalizations in our annual analysis was 3-4x times higher for influenza than COVID-19 among children <18 years, while the rates were 30 to 90% lower for influenza relative to COVID-19 among individuals aged 18+ years. In a US-based surveillance study focused on children, the COVID-19 hospitalization rate from October 2020–September 2021 was similar/lower to historical influenza seasons among children 0-11, but higher among adolescents aged 12–17.^27^

Our results need to be interpreted in the context of public health measures in place in BC throughout the first two years of the pandemic, diminishing the potential impact of COVID-19. These included testing within 24 hours for those who qualified, isolation and contact tracing for every case and contact, limits and restrictions on public and private gatherings, and mask wearing during periods of higher transmission. Businesses and schools adopted safety plans, and long-term care facilities were required to take preventive and outbreak measures at the order of Medical Health Officers. With the exception of a 9 week closure in March 2020, schools remained open for the entire duration of the pandemic. Nonetheless, extracurricular activities and gatherings were limited, reducing the contact rate among children and youth. In contrast, influenza hospitalizations represent pre-pandemic time periods without public health measures in place. Similar to other studies, we found that COVID-19 hospitalized patients were slightly more likely to be male^1,2,28^ (for all comparisons) and have a longer hospital stay.^2,28-31^ Overall median length of stay ranged from 6 (seasonal analysis – but no adjustment for primarily for COVID-19) to 8 days (annual analysis) for COVID-19 and ranged from 3 to 6 for influenza seasons, with differences more noticeable in patients >50 years of age. Length of stays for the annual analysis were close to other studies for COVID-19 in 2020 (median: 7^28^, 8^29^, and 9^30^) and influenza during different season (median=3^28^, 7^29^, and 5^30^).

This work fills an important gap in the literature comparing COVID-19 to influenza, as most evidence pertains to in-hospital populations rather than burden in the general population. Other notable strengths include the population-based capture of both influenza and COVID-19 hospitalizations and the ability to link to population-based COVID-19 immunization data. In addition, our comparative analysis alleviated potential biases by accounting for important factors, namely specific case ascertainment achieved by defining hospitalizations due primarily to COVID-19, COVID-19 vaccination status, and varying severity of different influenza strains.

However, the present analysis was hindered by the unavailability of individual-level influenza testing and immunization status and significance of COVID-19 as the reason for hospital admission. While the use of ICD codes to identify influenza hospitalizations is widely adopted in literature, some misclassification is expected. Yet, relying on positive influenza tests alone is not an ideal approach either: it underestimates the true number of influenza hospitalizations since testing practices differ by facility and not every patient with respiratory symptoms gets swabbed. In our validation analysis, the overall influenza estimates of hospitalization rates using ICD codes and test results were comparable with overlapping CIs. The time period for the annual analysis of COVID-19 was selected to capture mostly unvaccinated patients, however, in this time period not all variants of concern could not be included as this time period included rapidly evolving vaccination program.

In conclusion, our paper found that both COVID-19 and seasonal influenza resulted in “J” shaped hospitalization curves with the highest rates occurring at the extremities of age. Prior to widespread vaccination COVID-19 hospitalization rates were generally higher than those for influenza, with the exception of children and youth. Following vaccination, hospitalization rates for COVID-19 declined and became comparable to those attributable to influenza in spite of relaxing public health measures. These finding may have important implications for planning and preparation for future periods of influenza and SARS-CoV-2 co-circulation in fall 2022. Future studies are warranted to better understand the impact of COVID-19 relative to influenza in the context of no or little restrictions and/or co-circulation.

## Data Availability

The DAD and COVID-19 databases were made available through British Columbia COVID-19 Cohort (BCC19C), a public health surveillance platform integrating COVID-19 datasets (testing, cases, hospitalizations, vaccinations) with administrative data holdings for the BC population (e.g., medical visits, hospitalizations, emergency room visits, prescription drugs, chronic conditions, vital statistics). We are not permitted to share these data. BCC19C data are only available to researchers who request and meet the criteria for access.

## Contributors

KS, SS, and MZ were involved in the conception and design of the study. SS, MZ, and JW accessed and verified the data. SS was in charge of the study analysis. SS wrote the first draft. All authors were involved in the interpretation, critically reviewed one or multiple drafts of the manuscript, and approved the final version. KS and SS are the guarantors. The corresponding author attests that all listed authors meet authorship criteria and that no others meeting the criteria have been omitted.

## Ethical approval

This work was performed under BCCDC’s mandate to perform population health surveillance and falls under the Behavioural Research Ethics Board at the University of British Columbia (approval # H20-02097).

## Dissemination to participants and related patient and public communities

We will disseminate the findings to members of the public through press releases, knowledge translation products on institutional websites, as well as personal communication and social communication platforms.

## Transparency

The lead author and the senior author (the manuscript’s guarantors) affirm that this manuscript is an honest, accurate, and transparent account of the study being reported; that no important aspects of the study have been omitted; and that any discrepancies from the study as planned (and, if relevant, registered) have been explained.

## Acknowledgements

We acknowledge the assistance of the Provincial Health Services Authority, BC Ministry of Health and Regional Health Authority staff involved in data access, procurement, and management. We acknowledge BC Public Health Leadership table for their support and guidance. We acknowledge Dr. Mel Krajden, the medical director of the British Columbia Centre for Disease Control Public Health Laboratory, for his support in providing lab data for COVID-19 and influenza. We gratefully acknowledge the residents of British Columbia whose data are integrated in the British Columbia COVID-19 Cohort (BCC19C).

## Disclaimer

All inferences, opinions, and conclusions drawn in this manuscript are those of the authors, and do not reflect the opinions or policies of the Data Steward(s).

## Summary box

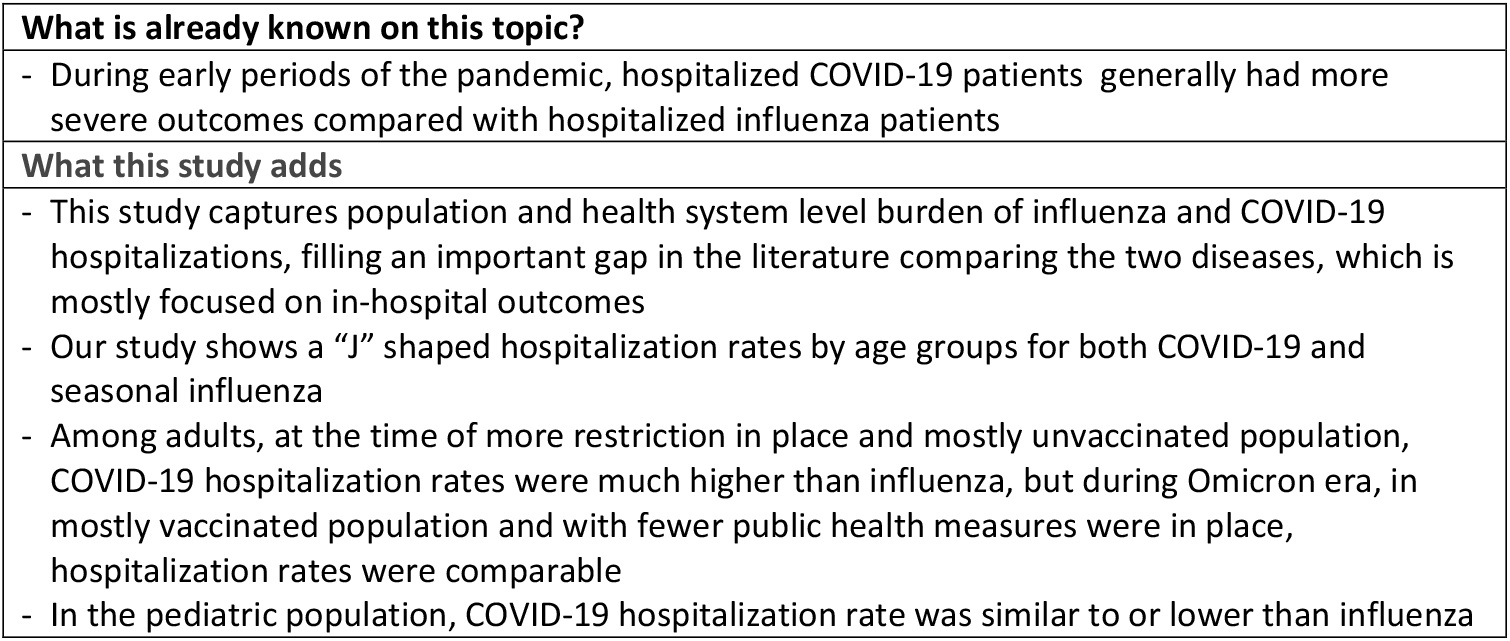

