## Supplemental Materials for "Comparison of influenza and COVID-19 hospitalizations in British Columbia, Canada: a population-based study"

*Table S1. Data Sources integrated within the British Columbia COVID-19 Cohort (BCC19C)*

| **British Columbia Centre for Disease Control (BCCDC), Provincial Heath Services Authority (PHSA) and Regional Health Authority data sources:** | **Data Date Ranges:** |
|---|---|
| - Integrated COVID-19 laboratory dataset (SARS-CoV2 tests from private/public labs) [S1] | Jan,2020-onward |
| - COVID-19 surveillance case data (information collected on all probable/confirmed cases as part of public health follow up by the regional health authorities in BC, including hospitalization data) [S2] | Jan,2020-onward |
| - Provincial COVID-19 Monitoring Solution (critical and non-critical care hospital census data, the PCMS is a daily hospital census of COVID-19 patients admitted to acute care facilities across BC and captures all hospitalizations following a positive COVID-19 test regardless of whether the hospitalization is for COVID-19) [S3] | Jan,2020-onward |
| - Provincial Immunizations Registry (COVID-19 vaccination data) [S4] | Dec,2020-onward |
| - Provincial Laboratory Information Solution (laboratory tests from private/public labs) [S5] | Jan,2020-onward |
| - Public Health Reporting Data warehouse (Influenza laboratory tests) [S6] | Jan,2008-onward |
| **Ministry of Health (MoH) Administrative Data Sources:** | |
| - Discharge Abstracts Database (DAD) (hospital discharge records) [S7] | 2008/9-onward |

*Table S2. Variables of interest of the study*

| Variables | Definition/Categories |
|---|---|
| Age | Age (years) was categorized into 0-4, 5-11, 12-17, 18-29, 30-39, 40-49, 50-59, 60-69, and 70+ years old |
| Sex | Sex was categorized into Male, Female, Unknown |
| Health authority of residence | British Columbia has five regional health authorities that deliver health services to meet the needs of the population within their respective geographic regions: Fraser, Vancouver Coastal, Vancouver Island, Interior, and Northern. Missing health authority was categorized as "Unknown". |
| Admission date | Date of hospital admission |
| Discharge date | Date of hospital discharge |
| Length of stay in hospital | Length of stay in the hospital was generated by subtracting discharge date from admission date within one episode of hospitalization |
| Vaccine status | Vaccine status was categorized into Fully vaccinated, Vaccinated with 1 dose (or partially vaccinated) and Unvaccinated. Fully vaccination was defined as receipt of two or more doses in which the second/third dose was received ≥14 days before case surveillance date. Case surveillance date is based on date of symptom onset. When symptom onset was unavailable, earliest laboratory date was used (collection or result date); if also unavailable, then public health report date was used. Partially vaccinated cases were those who became a case ≥21 days after their first dose and did not meet the criteria of fully vaccinated. Unvaccinated cases were those without any doses or those cases within <21 days after their first dose. Partially vaccinated cases were only included for the children 5-11 years old and excluded from 12+ cases due to small sample size. Children aged 0-4 were not eligible for COVID-19 vaccination. |

*Table S3: Demographic characteristics of hospitalized patients testing positive for COVID-19 during a year in 2020/21 and for influenza in 2009/10 (H1N1 pandemic), 2015/16 (severe for children), and 2016/17 (severe for adults)*

| **Characteristics** | **COVID-19 (n=3,398)** | **Influenza 2009/10 (n=1,483)** | **Influenza 2015/16 (n=1,032)** | **Influenza 2016/17 (n=1,979)** |
|---|---|---|---|---|
| | n (%) | n (%) | n (%) | n (%) |
| **Sex** | | | | |
| Female | 1462 (43) | 788 (53) | 519 (50) | 1037 (52) |
| Male | 1932 (57) | 695 (47) | 513 (50) | 941 (48) |
| Unknown | <5 | 0 (0) | 0 (0) | <5 |
| **Age, years** | | | | |
| Median (Q1-Q3) | 66 (52-78) | 40 (16-55) | 59 (33-75) | 77 (61-86) |
| **Age category, years** | | | | |
| 0-4 | 21 (1) | 196 (13) | 108 (10) | 67 (3) |
| 5-11 | 5 (0) | 130 (9) | 53 (5) | 29 (1) |
| 12-17 | 6 (0) | 68 (5) | 27 (3) | 23 (1) |
| 18-29 | 135 (4) | 195 (13) | 44 (4) | 39 (2) |
| 30-39 | 278 (8) | 143 (10) | 74 (7) | 69 (3) |
| 40-49 | 323 (10) | 217 (15) | 78 (8) | 73 (4) |
| 50-59 | 497 (15) | 243 (16) | 137 (13) | 168 (8) |
| 60-69 | 677 (20) | 123 (8) | 156 (15) | 258 (13) |
| 70+ | 1456 (43) | 168 (11) | 355 (34) | 1253 (63) |
| **Health authority** | | | | |
| Fraser | 1650 (49) | 460 (31) | 264 (26) | 439 (22) |
| Vancouver Coastal | 899 (26) | 279 (19) | 322 (31) | 636 (32) |
| Vancouver Island | 130 (4) | 228 (15) | 229 (22) | 601 (30) |
| Interior | 322 (9) | 351 (24) | 139 (13) | 224 (11) |
| Northern | 397 (12) | 151 (10) | 62 (6) | 59 (3) |
| Unknown | 0 (0) | 14 (1) | 16 (2) | 20 (1) |

Abbreviation: COVID-19=Coronavirus Disease 2019, n=number, Q=Quartile

Note: For the annual COVID-19 cohort we included all patients hospitalized for COVID-19 up to 14 days after or 2 days prior to specimen collection date from March 2020 to February 2021. For influenza, we selected three 12-month periods from September to August with distinct severity: 2009/10=H1N1 pandemic, 2015/16=mild influenza with higher severity in children, 2016/17=severe influenza with higher severity in adults

*Table S4: Median of length of stay (days) in hospital among patients testing positive for COVID-19 during a year in 2020/21 and patients hospitalized for influenza in 2009/10 (H1N1 pandemic), 2015/16 (severe for children), and 2016/17 (severe for adults) in British Columbia, Canada*

| **Age group, years** | **COVID_19 (n=3398)** | **Influenza 2009/10 (n=1483)** | **Influenza 2015/16 (n=1032)** | **Influenza 2016/17 (n=1979)** | **P-value[#]** |
|---|---|---|---|---|---|
| **Total** | | | | | |
| Median (Q1-Q3) | 8 (4-16) | 3 (2-6) | 5 (3-10) | 6 (3-12) | <0.001 |
| **0-4** | | | | | |
| Median (Q1-Q3) | 2 (2-3) | 2 (1-4) | 3 (2-5) | 3 (2-5) | 0.027 |
| **5-11** | | | | | |
| Median (Q1-Q3) | 2 (2-2) | 2 (2-5) | 3 (2-7) | 2 (1-5) | 0.043 |
| **12-17** | | | | | |
| Median (Q1-Q3) | 9.5 (2-20) | 2.5 (1.5-5) | 2 (1-4) | 3 (2-9) | 0.270 |
| **18-29** | | | | | |
| Median (Q1-Q3) | 4 (2-8) | 2 (2-4) | 3 (2-5) | 2 (1-3) | <0.001 |
| **30-39** | | | | | |
| Median (Q1-Q3) | 4 (2-7) | 3 (2-5) | 3 (2-8) | 3 (2-5) | <0.001 |
| **40-49** | | | | | |
| Median (Q1-Q3) | 6 (3-11) | 4 (2-7) | 4 (3-9) | 4 (3-7) | <0.001 |
| **50-59** | | | | | |
| Median (Q1-Q3) | 7 (4-12) | 5 (2-8) | 7 (3-13) | 4 (2-8) | <0.001 |
| **60-69** | | | | | |
| Median (Q1-Q3) | 9 (5-17) | 5 (2-9) | 6 (3-10) | 5 (3-11) | <0.001 |
| **70+** | | | | | |
| Median (Q1-Q3) | 10 (5-19) | 5 (3-10.5) | 7 (4-12) | 7 (4-14) | <0.001 |

Abbreviation: COVID-19= Coronavirus Disease 2019, n=number, Q=Quartile

Note: For the annual COVID-19 cohort we included all patients hospitalized for COVID-19 up to 14 days after or 2 days prior to specimen collection date from March 2020 to February 2021. For influenza, we selected three 12-month periods from September to August with distinct severity: 2009/10=H1N1 pandemic, 2015/16=mild influenza with higher severity in children, 2016/17=severe influenza with higher severity in adults

#Kruskal–Wallis one-way analysis of variance

*Table S5: Validation of ICD-10 algorithms against lab-confirmed hospitalization to identify hospitalized individuals with seasonal influenza infection residing in Vancouver Coastal Health region of British Columbia, Canada*

| Characteristics | TP | FP | FN | TN | Sensitivity (95% CI) | Specificity (95% CI) | PPV (95%CI) | NPV (95%CI) |
|---|---|---|---|---|---|---|---|---|
| **Influenza 2009/10** | 142 | 114 | 20 | 406 | 88 (83, 93) | 78 (75, 82) | 55 (49, 62) | 95 (93, 97) |
| **Influenza 2015/16** | 154 | 21 | 62 | 652 | 71 (65, 77) | 97 (95, 98) | 88 (82, 92) | 91 (89, 93) |
| **Influenza 2016/17** | 361 | 64 | 121 | 1094 | 75 (71, 79) | 94 (93, 96) | 85 (82, 88) | 90 (88, 92) |

Abbreviation: CI=Confidence Interval, FN=False Negative, FP=False Positive, ICD-10=International Classification of Disease 10$^{th}$ edition, NPV=Negative Predictive Value, PPV=Positive Predictive Value, TN=True Negative, TP=True Positive
Note: We selected the 8-week peak in each influenza season.

A

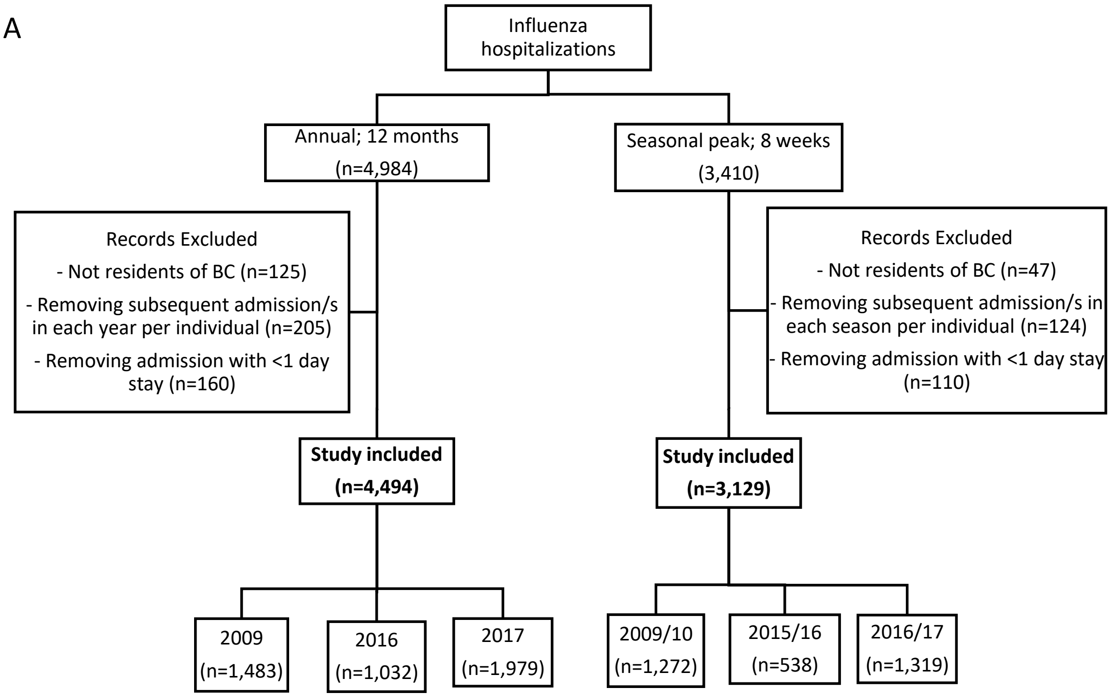


B

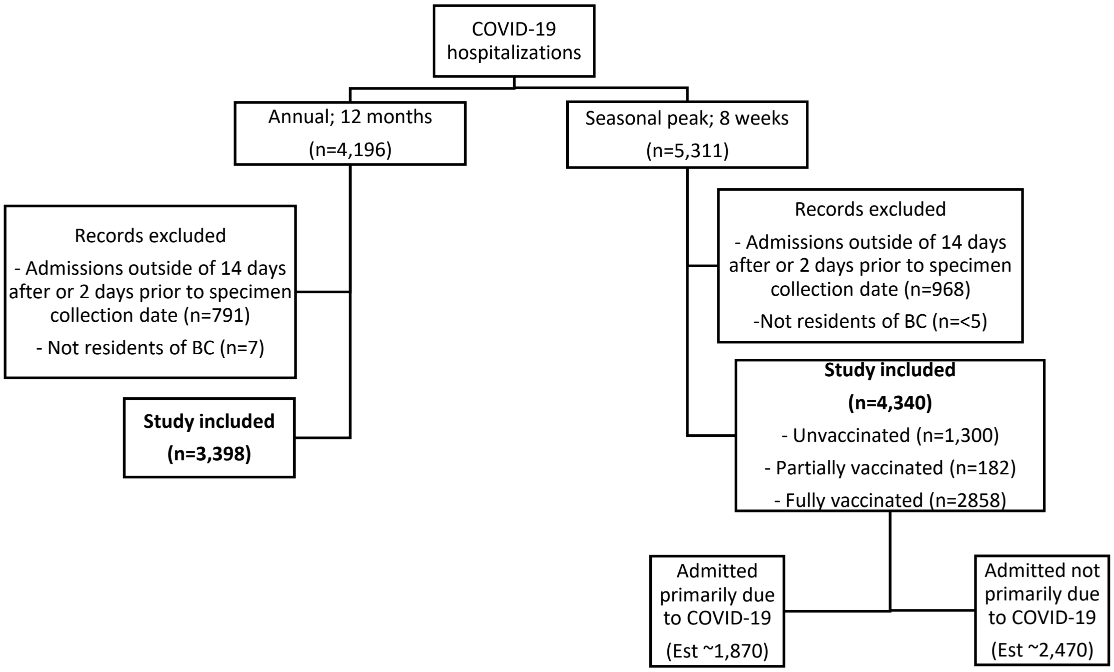


*Figure S1: Annual and seasonal peak study cohorts for influenza and COVID-19 hospitalization.*

*Note:* A) In annual influenza we selected three years from September to August with distinct severity: 2009/10=H1N1 pandemic, 2015/16=mild influenza with higher severity in children, 2016/17=severe influenza with higher severity in adults. For influenza seasonal analysis we selected the 8-week peak from each season. B) For the annual COVID-19 cohort we included all patients hospitalized for COVID-19 up to 14 days after or 2 days prior to specimen collection date from March 2020 to February 2021. For the peak COVID-19 cohort we included all patients hospitalized for COVID-19 up to 14 days after or 2 days prior to specimen collection date during the first 8 weeks of 2022 when Omicron was dominant and >90% of adults in British Columbia were vaccinated with at least 2 doses. We applied the following estimates for proportion of hospitalizations likely primarily due to COVID-19:

unvaccinated <50 years old (50%), unvaccinated ≥50 years old (70%), vaccinated <50 years old (20%) and vaccinated ≥50 years old (40%). Numbers were rounded to zero decimal point.

Figure S2: Hospitalization rates in patients testing positive for COVID−19 during the peak of Omicron variant (week 1−8 of 2022) compared with hospitalized patients for influenza in the peak of 2009/10 (H1N1 pandemic), 2015/16 (severe for children), and 2016/17 (severe for adults) season, by age group and COVID−19 vaccine status, British Columbia, Canada

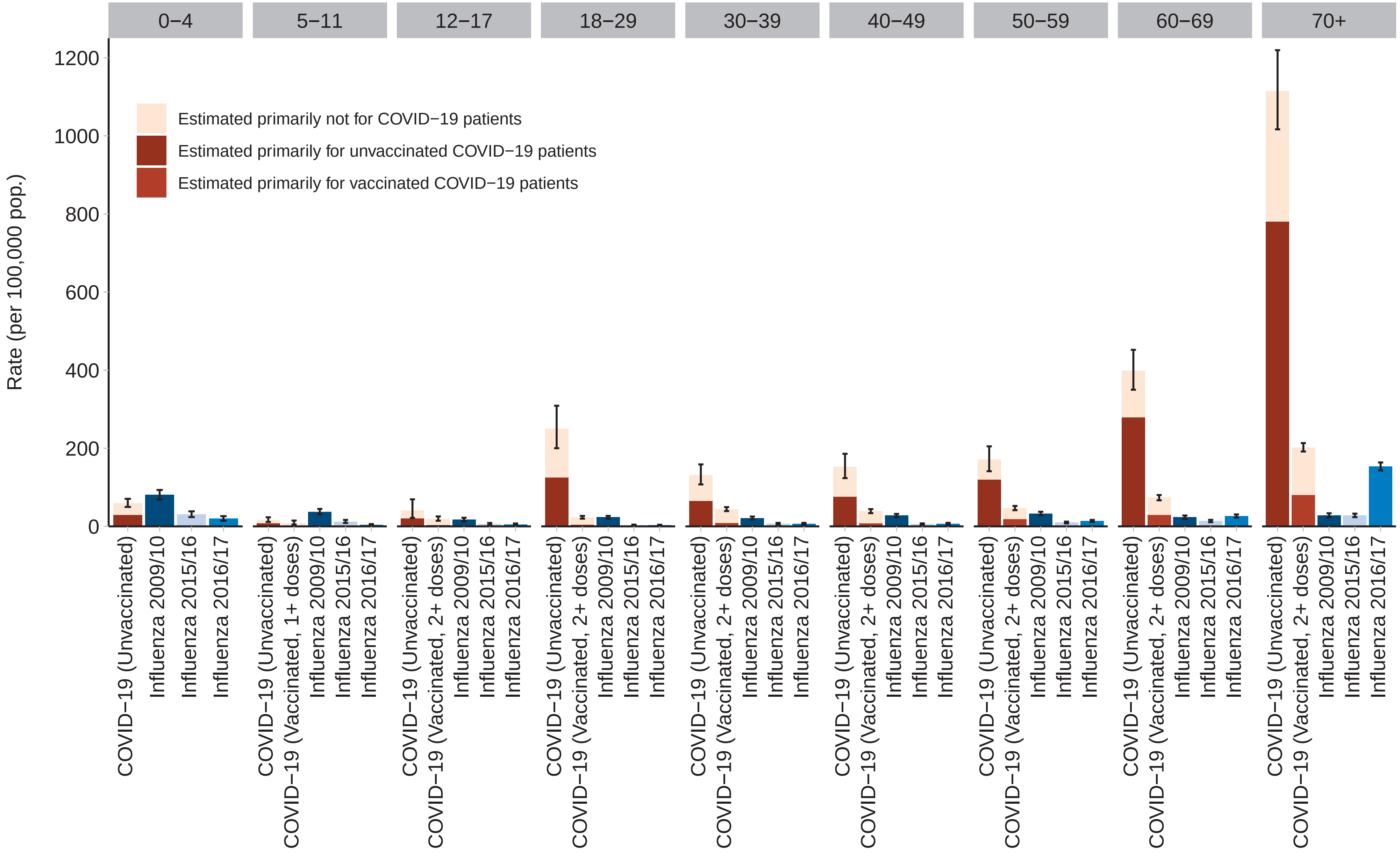


Note: For the peak COVID−19 cohort we included all patients hospitalized for COVID−19 up to 14 days after or 2 days prior to specimen collection date during the first 8 weeks of 2022 when Omicron was dominant and >90% of adults in British Columbia were vaccinated with at least 2 doses. We selected the 8−week peak of three seasons with distinct severity for influenza: 2009/10=H1N1 pandemic, 2015/16=mild with higher severity in children, 2016/17=severe with higher severity in adults. We applied the following estimates for proportion of hospitalizations likely primarily due to COVID−19: unvaccinated <50 years old (50%), unvaccinated >=50 years old (70%), vaccinated <50 years old (20%) and vaccinated >=50 years old (40%).

Figure S3: Seasonal lab−confirmed versus ICD code−identified (FLU2 algorithm) influenza hospitalization rates during the peak (8 weeks) of 2009/10 (H1N1 pandemic), 2015/16 (severe for children), and 2016/17 (severe for adults), by age group among the residents of Vancouver Coastal Health, British Columbia, Canada

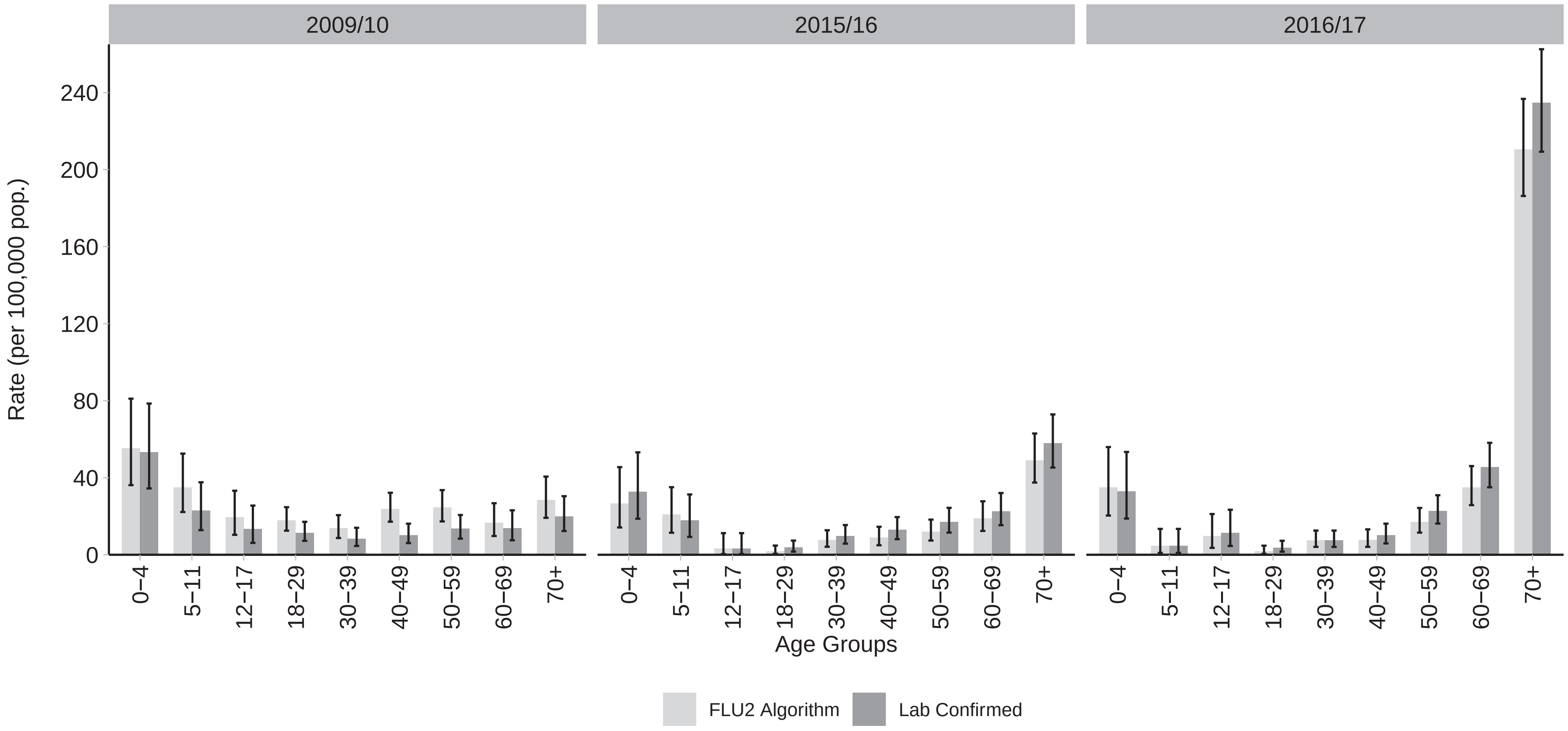


Note: We selected the 8−week peak of three seasons with distinct severity for influenza: 2009/10=H1N1 pandemic, 2015/16=mild with higher severity in children, 2016/17=severe with higher severity in adults.
